# Serum Oxidative Stress Markers in Women with Uterine Fibroids in Lagos, Nigeria

**DOI:** 10.1101/2021.07.07.21260056

**Authors:** Adaiah Priscillia Soibi-Harry, Christian Chigozie Makwe, Ayodeji Ayotunde Oluwole, Sunusi Rimi Garba, Abisoye Towuromola Ajayi, Roosevelt Anyanwu, Rose Ihuoma Anorlu

## Abstract

**Background:** Uterine fibroid is the most common benign gynaecological tumour in women of reproductive age group, with significant impact on the quality of life, economy, morbidity and sometimes mortality of affected women. Black women of reproductive age group are more likely to develop uterine fibroids. Although available evidence suggests racial and genetic predisposition to the aetiology of uterine fibroid, oxidative stress has been implicated in the onset and progression of uterine fibroids. Epidemiological and experimental studies have suggested that oxidative stress may play an important role in the pathogenesis of gynaecological diseases including uterine fibroids.

**Aim and Objectives:** This study aims to assess the serum levels of antioxidants (catalase (CAT), superoxide dismutase (SOD), glutathione peroxidase (GPx)) and oxidants (protein carbonyl (PC), advanced oxidation protein products (AOPP)) in women with uterine fibroids and to identify any association between the size of uterine fibroids and serum levels of the analysed antioxidants and oxidants.

**Methods:** Forty-four women with ultrasound diagnosis of uterine fibroids and 44 women without uterine fibroids were recruited from a University Teaching Hospital. Blood samples were obtained and analysed for serum levels of selected antioxidants (CAT, SOD, GPx) and oxidants (PC, AOPP). Pelvic ultrasonography was performed on all study participants.

**Results:** The median serum levels of antioxidants: CAT (2.20 vs 4.32 ng/ml; p < 0.001); SOD (285.54 vs 380.96 ng/ml; *p* < 0.001) and GPx (9.67 vs 11.26 µU/ml; *p* < 0.001) were significantly lower in women with uterine fibroids. The median serum levels of oxidants: PC (162.08 vs 142.36 ng/ml; *p* = 0.04); and AOPP (22.42 vs 13.94 ng/ml; *p* < 0.001) were significantly higher in women with uterine fibroids. There was a strong negative correlation between serum levels of AOPP and SOD (r = -0.95; *p* < 0.001) in women with uterine fibroids. The maximum diameter of fibroids showed a significant positive correlation with AOPP (r = 1.000; *p* < 0.001) and a negative correlation with SOD (r = -1.000; *p* < 0.001).

**Conclusion:** Women with uterine fibroids had lower levels of antioxidants and higher levels of oxidants. In women with uterine fibroids, AOPP negatively correlated with SOD. There was a positive correlation between fibroid size and AOPP and a negative correlation between fibroid size and SOD. The findings of this study suggests that AOPP and SOD may play an important role in uterine fibroids.

## Introduction

Uterine fibroids are a disease of public health importance as the most common benign tumour in women of reproductive age, with significant impact on their quality of life. It places a huge demand on scarce health care resources, and is the single most common reason for hysterectomy globally.^1,2^ About 70% of Caucasian and 80% of black women, will be diagnosed with uterine fibroid at least once in their lifetime.^3^ Black women are three times more likely to have uterine fibroids with an earlier onset of disease, more severe symptoms, a lower quality of life and more missed days at work.^4^ It is unclear what the true incidence and prevalence of uterine fibroid in Nigerian women is, however, estimates of 19.7% to 31% have been reported in facility-based studies.^5,6^

The pathogenesis of uterine fibroids is still evolving with several proposed aetiologies and associations.^1^ Uterine fibroids are characterized by an increase in smooth muscle cell proliferation and large deposits of extracellular matrix proteins.^1,7^ Reactive oxygen species (ROS) seem to be involved in the signaling pathway of several growth factors, cytokines, and vasoactive agents that stimulate proliferation of a variety of cell types including uterine fibroids.^8^ ROS are oxygen containing chemicals produced by living organisms in the course of normal cellular metabolism.^9^ At moderate concentrations, ROS function in cell signaling and homeostasis; but at high concentrations, they modify cell components such as lipids, proteins, and deoxyribonucleic acid (DNA).^10^

Oxidative stress occurs when there is a shift in balance between oxidants and antioxidant in favour of oxidants, with consequent overriding of the scavenging capacity by antioxidants, either due to the reduced availability of antioxidants or excessive generation of ROS. This phenomenon can cause DNA modification by degrading bases, breaking DNA and modifying purines and pyrimidines, thus resulting in mutations, deletions, translocations and cross-linking with proteins.^11^ These modifications have been linked to many pathological conditions, such as cancer, profibrotic gynaecologic diseases like uterine fibroids, neurological disorders, atherosclerosis, and so on.^10^ Segars et al. proposed that the large amounts of extracellular matrix which make up uterine fibroids may have undergone oxidative stress and resulted in an imbalance between connective tissue production and degradation, accumulation of myofibroblasts, and deposition of newly formed extracellular matrix component.^12^

Advanced oxidative protein products (AOPP) and protein carbonyl (PC) are recognised oxidative stress biomarkers that result from the oxidative modification of proteins. AOPP are formed when chloramines and hypochlorous acid react with plasma proteins.^13^ PC results from either oxidation of amino acids residues (cysteine, methionine, lysine, arginine), cleavage of protein backbone, adduction of reactive aldehydes or the amidation pathway. Enzymatic antioxidants such as catalase (CAT), superoxide dismutase (SOD), glutathione peroxidase (GPx) and glutathione reductase (GR) directly destroys reactive oxygen species.^11^

There is increasing interest and research on the role of oxidative stress in the pathogenesis of uterine fibroids.^14–17^ In a study by Santuli et al., they found significantly higher levels of AOPP and PC in the serum of women with uterine fibroids.^18^ It has been suggested that certain antioxidant supplements may be useful in the management of some gynaecologic diseases.^19–22^ There is paucity of data on the relationship between uterine fibroids and oxidative stress among black women in Sub-Saharan Africa, including Nigeria. This study assessed and compared the levels of antioxidants and oxidants among reproductive-aged women with and without uterine fibroids. The findings from this study will help bridge the knowledge gap on the possible role of oxidative stress in uterine fibroids.

## Materials and Method

This was a cross sectional study carried out at the Out-patient Clinic of Lagos University Teaching Hospital (LUTH) over a period of six months from July 2019 to January 2020, after obtaining ethical approval from the LUTH Health Research Ethic Committee (ADM/DCST/HREC/APP/2722). Eighty-eight eligible women, aged 20 -45 years were recruited into the study after obtaining informed written consent from each participant. An anonymous structured questionnaire was used to obtain relevant information by face-to-face interview. Information sought included: socio-demographic data, medical history, surgical history, obstetric history, analgesic use, anthropometry, menstrual history, history of dysmenorrhea, deep dyspareunia, non-cyclic chronic pelvic pain, gastrointestinal and lower urinary tract symptoms, history of abnormal uterine bleeding and history of uterine fibroids in first degree relatives.

Excluded from the study were pregnant and lactating women, current users of hormonal contraceptives, women who smoked cigarette or had chronic medical conditions such as endometriosis, cancer, hypertension, hypercholesterolemia, cardiovascular disease, diabetes, renal disease, tuberculosis, human immunodeficiency virus and Hepatitis B or C. These conditions were considered potential confounders for altered serum levels of antioxidants and oxidants.

Pelvic ultrasonography was performed on all participants using a 3D ultrasound unit TUS-X100S (Toshiba Diagnostic Ultrasound Systems Ltd., Japan) with a transabdominal (5.0 MHz) and/or transvaginal (7.0 MHz) ultrasound probes as appropriate. Pelvic ultrasonography was interpreted in real-time. Participants without uterine fibroids were age matched (within ± 2 years) to those with uterine fibroids and assigned to the no fibroid group (n = 44). For participants with sonographic evidence of uterine fibroid, the number, location and size of fibroids were documented. Measurement of each fibroid was performed in sagittal and axial views and the maximum diameter was retained for the study. The participants with sonographic evidence of uterine fibroids were assigned to the fibroid group (n = 44)

### Sample Collection

Specimen bottles were labelled with the unique identification number of each participant and 5ml of venous blood was obtained with a peripheral venous catheter into a plain vacutainer. The sample was allowed to clot for 20 minutes at room temperature, centrifuged using Eppendorf 5415C centrifuge (Eppendorf AG, Germany) at 2500 rpm for 20 minutes. The serum supernatant was removed with a sterile Pasteur pipette without disturbing the white buffer layer and equal volumes were aliquoted into two cryogenic vials and frozen at -80°C. The stored samples were brought out of the freezer and allowed to thaw on the work bench and all assays were performed at room temperature as a batch.

### Assay of antioxidants and oxidants

The following kits were used for the assays: Human AOPP Elisa Kit, E1266Hu; Human PC Elisa Kit, E1426Hu; Human Extracellular [SOD] [Cu-Zn] Elisa Kit, E4236Hu; Human CAT Elisa Kit E3053Hu; Human GPx-1 Elisa Kit E3921Hu; and Human CRP Elisa Kit E1805Hu (Bioassay Technology Laboratory, Shanghai, China). The values for the limit of detection (LOD) of the different assays (AOPP, PC, SOD, CAT, GPx-1, and CRP) were 0.1 ng/mL, 2 ng/mL, 0.5 ng/mL, 2 ng/mL, 3 µU/mL, and 0.3mg/L respectively. The intra-and inter-assay variations of all the assays were similar (<8 and <10%, respectively). Automated pipettes were recalibrated before use and commercially prepared control sera were used simultaneously for all analysis. The manufacturer’s instructions were strictly followed

### Statistical Analysis

Data was analysed using the IBM Statistical Package for Social Sciences (SPSS) Statistics for Windows, Version 25.0 [Armonk, NY: IBM Corp]. The data was summarized and presented as tables, figures and graphs. Test of normality was performed using Shapiro-Wilk test. The normally distributed continuous variables were presented as mean ± standard deviation, while non-parametric variables were presented as median ± interquartile range. Mann-Whitney U Test was used to compare the serum levels of antioxidants and oxidants. Serum oxidative stress parameters were represented as boxes and whisker plots. Student’s T-test was used to compare the normally distributed continuous variables. Chi square test was used for comparison of categorical variables. Simple linear regression analysis was done to assess the relationship between antioxidants and oxidants in the serum of women with and without uterine fibroids. Multiple regression analysis was used to examine the influence of AOPP, PC, SOD, CAT and GPx on the maximum diameter of fibroids. *P* value less than 0.05 was considered statistically significant.

## Results

A total of 88 women who met the selection criteria were recruited for the study and assigned to two groups; 44 participants diagnosed with uterine fibroids and 44 participants without uterine fibroids based on pelvic ultrasonography. There was 100% agreement between the sonologists (ASH and ATA) on the ultrasonographic diagnosis of uterine fibroids and the assignment of study groups. Data were complete and all study participants were included in the statistical analysis.

The sociodemographic and baseline characteristics of the two groups are presented on Table1. There was no statistically significant difference in age, parity, level of education, marital status, employment status and socio-economic status between the two groups.

**TABLE 1.** SOCIODEMOGRAPHIC AND BASELINE CHARACTERISTICS.

| VARIABLES | FIBROID<br>(N = 44) | NO FIBROID<br>(N = 44) | $\chi^2$ | *p* -value |
| --- | --- | --- | --- | --- |
|  | Mean (SD) | Mean (SD) |  |  |
| Age (years) | 38.3 (6.5) | 37.9 (7.5) | 1.55* | 0.132 |
| BMI(Kg/m<sup>2</sup>) | 26.88 (5.18) | 26.14 (4.38) | 0.76* | 0.470 |
|  | n (%) | n (%) |  |  |
| Parity |  |  |  |  |
| **0** | 28 (63.6) | 27 (61.4) | 0.35 | 0.837 |
| **1** | 6 (13.7) | 8 (18.2) |  |  |
| **≥ 2** | 10 (22.7) | 9 (20.4) |  |  |
| Ethnicity |  |  |  |  |
| **Igbo** | 11 (25) | 22 (50) | 7.84 | 0.054 |
| **Yoruba** | 22 (50) | 18 (40.9) |  |  |
| **Hausa** | 1 (2.3) | 1 (2.3) |  |  |
| **Others** | 10 (22.7) | 3 (6.8) |  |  |
| Level of education |  |  |  |  |
| **Primary** | 1 (2.3) | 1 (2.3) | 0.00 | 1.000 |
| **Secondary** | 7 (15.9) | 7 (15.9) |  |  |
| **Tertiary** | 36 (81.8) | 36 (81.8) |  |  |
| Employment status |  |  |  |  |
| **Employed** | 36 (81.8) | 32 (72.7) | 2.92 | 0.234 |
| **Unemployed** | 7 (15.9) | 7 (15.9) |  |  |
| **Student** | 1 (2.3) | 5 (11.4) |  |  |
| Marital Status |  |  |  |  |
| **Married** | 26 (59.1) | 26 (59.1) | 0.00 | 1.000 |
| **Single** | 15 (34.1) | 15 (34.1) |  |  |
| **Separated** | 3 (6.8) | 3 (6.8) |  |  |
| Socioeconomic status |  |  |  |  |
| **Semi-skilled** | 15 (34.1) | 20 (45.5) | 1.41 | 0.493 |
| **Skilled professional** | 24 (54.5) | 21 (47.7) |  |  |
| **Unskilled** | 5 (11.4) | 3 (6.8) |  |  |

SD- Standard deviation, \*Student's T-test

Figure 1 shows significantly lower median serum levels of antioxidants in women with uterine fibroids when compared to women without fibroids: CAT (2.20 vs 4.32 ng/ml; p < 0.001); SOD (285.54 vs 380.96 ng/ml; *p* < 0.001) and GPx (9.67 vs 11.26 µU/ml; *p* < 0.001). The median serum levels of oxidants were significantly higher in women with uterine fibroids: PC (162.08 vs 142.36 ng/ml; *p* = 0.04); and AOPP (22.42 vs 13.94 ng/ml; *p* < 0.001). Table 2 shows the correlation of oxidants and antioxidants in women with and without uterine fibroids. There was a statistically significant negative correlation between SOD and AOPP (r = -0.95; p < 0.001) in women with uterine fibroids.

**TABLE 2.** CORRELATION OF ANTIOXIDANTS AND OXIDANTS IN WOMEN WITH AND WITHOUT UTERINE FIBROIDS.

| Spearman Rank Correlation |  |  |  |  |  |  |
| --- | --- | --- | --- | --- | --- | --- |
| Coefficient (r) (p value) |  |  |  |  |  |  |
| Variables | CAT |  | SOD |  | GPx |  |
|  | Fibroid | No fibroid | Fibroid | No fibroid | Fibroid | No fibroid |
|  | (n = 44) | (n = 44) | (n = 44) | (n = 44) | (n = 44) | (n = 44) |
| **PC** | -0.09<br>(0.57) | 0.07 (0.68) | - 0.11<br>(0.46) | 0.01<br>(0.95) | 0.13 (0.39) | 0.20 (0.20) |
| **AOPP** | 0.02<br>(0.89) | -0.01 (0.94) | - 0.95<br>(<0.001) | 0.03<br>(0.85) | 0.26 (0.08) | -0.10(0.51) |

There was a positive correlation between the maximum diameter of fibroids and serum levels of AOPP (r = 1.00; p < 0.001) and a negative correlation between the maximum diameter of fibroids and serum levels of SOD (r = -1.00; p < 0.001). An extra 1 ng/ml increase in serum level of AOPP, was associated with a 3.77 mm, [95% CI: 3.27-4.26] increase in the diameter of uterine fibroids, while a 1 ng/ml increase in serum level of SOD, was associated with a 1.10mm, [95% CI: -1.24 -0.97] decrease in the diameter of uterine fibroids. (Table 3)

**TABLE 3.** MULTIPLE REGRESSION ANALYSIS OF OXIDATIVE STRESS MARKERS AND MAXIMUM DIAMETER OF FIBROIDS (N = 44)

| Variables | Correlation<br>with maximum<br>diameter of<br>uterine fibroid<br>(r) | Multiple regression weights |  | t | Sig. | 95% CI for B |  |
| --- | --- | --- | --- | --- | --- | --- | --- |
|  |  | B | beta |  |  | Lower<br>Bound | Upper<br>Bound |
| **PC (ng/ml)** | 0.11 | - 0.01 | - 0.02 | -0.10 | 0.92 | - 0.28 | 0.25 |
| **AOPP**<br>**(ng/ml)** | 1.00 | 3.77 | - 0.04 | 15.43 | < 0.001 | 3.27 | 4.26 |
| **CAT**<br>**(ng/ml)** | 0.02 | - 5.79 | - 0.06 | - 0.37 | 0.72 | -37.79 | 26.22 |
| **SOD**<br>**(ng/ml)** | -1.00 | -1.10 | - 0.93 | -16.39 | < 0.001 | -1.24 | - 0.97 |
| **GPx**<br>**(μU/ml)** | 0.26 | 7.41 | 0.28 | 1.91 | 0.06 | - 0.43 | 15.26 |

## Discussion

This study found statistically significant reduced levels of antioxidants (CAT, SOD and GPx) and increased levels of oxidants (PC and AOPP) in the serum of women with uterine fibroids compared to women without uterine fibroids. Serum levels of CAT, SOD and GPx, which are major enzymatic antioxidants that function as scavengers of reactive oxygen species were significantly lower in the women with uterine fibroids compared to those without uterine fibroids. This is consistent with the findings of Oyeyemi, of reduced activity of antioxidants (SOD, CAT and GPx) in the serum of women with uterine fibroids, but contrasting with the findings of Pejic et al., who did not find any statistical difference between serum levels of CAT in women with fibroids compared to those without fibroids.^23,24^ This observation may be due to the difference in the race of the study population. Black race has been identified as an independent factor for increased oxidative stress as well as uterine fibroids.^4,25^ It is possible that the predisposition of black women to higher levels of oxidative stress, may have resulted in the consumption of CAT within the cells to achieve redox balance with subsequent reduction in CAT levels in the serum of women in this study.

This study found increased levels of the oxidants (AOPP and PC) which is in agreement with the findings of Santuli et al.^18^ This may be because of the similarities in the study design. The elevated levels of oxidants in women with uterine fibroids is not related to acute inflammatory process or reaction because the levels of C -reactive protein (CRP) in both groups of women were within the normal range for CRP, < 10 mg/L.^26^ In addition, there was no statistically significant difference in the serum levels of CRP between the women with uterine fibroids and women without uterine fibroids. This implies that the recruitment strategy for selection of eligible participants into the study eliminated the potential enrolment of women with conditions associated with elevated CRP or acute inflammatory responses.

The relationship and interaction between antioxidants and oxidants in women with and without uterine fibroids were explored. There was a statistically significant negative correlation between AOPP and SOD in women with uterine fibroids and no correlation found in those without uterine fibroids. This is in agreement with the findings of Pejic et al., where they found a significant negative correlation between oxidants and antioxidants.^24^ Although the sample size used in their study was relatively small, this study has corroborated their finding that indeed there is an interplay between antioxidants and oxidants in women with uterine fibroids. The reduced levels of antioxidants in response to the elevated levels of oxidants seen in this study, may be a compensatory activity within the cells, where antioxidants are used up to maintain redox balance. This gives credence to the hypothesis of oxidative damage in uterine fibroids.^27^

The negative correlation between AOPP and SOD suggests that the redox activity in uterine fibroids, may be as a result of the interplay between AOPP and SOD. It is also possible that SOD, an enzymatic antioxidant scavenger, may have been used up while trying to balance the effect of the elevated oxidant, AOPP. This mimics the normal physiologic role of antioxidants, in maintaining redox balance within the cells.

The maximum diameter of fibroids showed a statistically significant positive correlation with the oxidant AOPP and negative correlation with the antioxidant SOD. This is in agreement with the findings of Vural et al. and Santuli et al., who found that fibroid size correlated positively with serum concentration of oxidants and negatively with antioxidants.^16,18^ The findings from this study suggest a relationship between oxidative stress and fibroid size. However, whether the observed finding is a cause or an effect of uterine fibroids, remains to be elucidated. More so, the normal range for serum levels of oxidative stress parameters are still been debated.^28^ From this study we may infer that larger fibroid sizes were associated with higher serum levels of the oxidant (AOPP) and lower serum levels of the antioxidant (SOD).

The strength of this study involves the fact that it was carried out in amongst black African women, the laboratory method employed for the determination of levels of oxidative stress markers in this study was the ELISA technique which is more accurate than previously employed methods as it measured the exact concentrations instead of activity of the analytes. Participants were age matched, and those with infectious and chronic diseases were excluded from the study to reduce confounders. Levels of CRP was determined for the two groups of women in order to exclude acute inflammation.

Some potential limitations which should be considered when interpreting this study include the fact that reactive oxygen species may be produced by other disease conditions that had not been previously diagnosed and the participants were not aware of such condition. In addition, this was a hospital-based study and may not be a true representation of the general population of women with uterine fibroids. This study was carried out on participants with ultrasonographic diagnosis of uterine fibroids and there was no histopathological confirmation of the diagnosis.

## Conclusion

Women with uterine fibroids had lower levels of antioxidants and higher levels of oxidants when compared to women without uterine fibroids. In women with uterine fibroids, AOPP negatively correlated with SOD. There was a positive correlation between fibroid size and AOPP and a negative correlation between fibroid size and SOD. The findings of this study suggests that AOPP and SOD may play an important role in uterine fibroids.

## Data Availability

Data will be available after obtaining permission from my institution

## Acknowledgments

The researchers wish to acknowledge Mr. Taiwo Adesina, College of Medicine, University of Lagos, Nigeria.

## References

1. Stewart EA, Laughlin-Tommaso SK, Catherino WH, Lalitkumar S, Gupta D, Vollenhoven B.. Uterine fibroids. Nature Reviews Disease Primers. 2016;2: 16043.

2. Whiteman MK, Hillis SD, Jamieson DJ, Morrow B, Podgornik MN, Brett KM, et al. Inpatient hysterectomy surveillance in the United States, 2000-2004. Am J Obstet Gynecol. 2008;198: 34.e1-7.

3. Baird DD, Dunson DB, Hill MC, Cousins D, Schectman JM. High cumulative incidence of uterine leiomyoma in black and white women: ultrasound evidence. Am J Obstet Gynecol. 2003;188: 100–107.

4. Stewart EA, Nicholson WK, Bradley L, Borah BJ. The burden of uterine fibroids for African-American women: results of a national survey. Journal of women’s health. 2013;22: 807–816.

5. Elugwaraonu O, Okojie AI, Okhia O, Oyadoghan GP. The Incidence of Uterine Fibroid Among Reproductive Age Women: A Five-Year Review of Cases at Isth, Irrua, Edo, Nigeria. International Journal of Basic, Applied and Innovative Research. 20131;2: 55–60.

6. Oluwole AA, Owie E, Babah OA, Afolabi BB, Oye-Adeniran BA. Epidemiology of Uterine Leiomyomata at the Lagos University Teaching Hospital, Idi-Araba, Lagos. Nigerian Hospital Practice. 2015;15: 14–20.

7. Grudzien MM, Low P, Manning PC, Arredondo M, Belton RJ, Nowak RA. The Antifibrotic Drug Halofuginone Inhibits Proliferation and Collagen Production by Human Leiomyoma and Myometrial Smooth Muscle Cells. Fertil Steril. 2010;93: 1290–1298.

8. Mesquita FS, Dyer SN, Heinrich DA, Bulun SE, Marsh EE, Nowak RA. Reactive Oxygen Species Mediate Mitogenic Growth Factor Signaling Pathways in Human Leiomyoma Smooth Muscle Cells. Biol Reprod. 2010;82: 341–351.

9. Liguori I, Russo G, Curcio F, Bulli G, Aran L, Della-Morte D, Gargiulo G, Testa G, Cacciatore F, Bonaduce D, Abete P. Oxidative stress, aging, and diseases. Clin Interv Aging. 2018;13: 757–772.

10. Elsayed Azab a, A Adwas A, Ibrahim Elsayed AS, A Adwas A, Ibrahim Elsayed AS, Quwaydir FA. Oxidative stress and antioxidant mechanisms in human body. JABB. 2019;6: 43–47.

11. Birben E, Sahiner UM, Sackesen C, Erzurum S, Kalayci O. Oxidative Stress and Antioxidant Defense. World Allergy Organ J. 2012;5: 9–19.

12. Segars JH, Parrott EC, Nagel JD, Guo XC, Gao X, Birnbaum LS, et al. Proceedings from the Third National Institutes of Health International Congress on Advances in Uterine Leiomyoma Research: comprehensive review, conference summary and future recommendations. Hum Reprod Update. 2014;20: 309–333.

13. Selmeci L, “Advanced oxidation protein products (AOPP): Novel uremic toxins, or components of the non-enzymatic antioxidant system of the plasma proteomeã” Free Radical Research. 2011;45: 1115–1123.

14. Fletcher NM, Saed MG, Abuanzeh S, Abu-Soud HM, Al-Hendy A, Diamond MP, et al. Nicotinamide Adenine Dinucleotide Phosphate Oxidase Is Differentially Regulated in Normal Myometrium Versus Leiomyoma. Reprod Sci. 2014;21:1145–1152.

15. Foksinski M, Kotzbach R, Szymanski W, Olinski R. The level of typical biomarker of oxidative stress 8-hydroxy-2’-deoxyguanosine is higher in uterine myomas than in control tissues and correlates with the size of the tumor. Free Radic Biol Med. 2000;29: 597–601.

16. Vural M, Camuzcuoglu H, Toy H, Camuzcuoglu A, Aksoy N. Oxidative stress and prolidase activity in women with uterine fibroids. J Obstet Gynaecol. 2012;32: 68–72.

17. Pejić S, Todorović A, Stojiljković V, Gavrilović L, Popović N, Pajović SB. Antioxidant status in women with uterine leiomyoma: relation with sex hormones. Anais da Academia Brasileira de Ciências. 2015;87: 1771–1782.

18. Santulli P, Borghese B, Lemaréchal H, Leconte M, Millischer AE, Batteux F, Chapron C, Borderie D. Increased serum oxidative stress markers in women with uterine leiomyoma. PloS one. 2013;8: e72069.

19. Zhang D, Al-Hendy M, Richard-Davis G, Montgomery-Rice V, Rajaratnam V, Al-Hendy A. Antiproliferative and Proapoptotic Effects of Epigallocatechin Gallate on Human Leiomyoma Cells. Fertil Steril. 2010;94: 1887–1893.

20. Sahin K, Ozercan R, Onderci M, Sahin N, Khachik F, Seren S, et al. Dietary tomato powder supplementation in the prevention of leiomyoma of the oviduct in the Japanese quail. Nutr Cancer. 2007;59: 70–75.

21. Santanam N, Kavtaradze N, Murphy A, Dominguez C, Parthasarathy S. Antioxidant Supplementation Reduces Endometriosis Related Pelvic Pain in Humans. Transl Res. 2013;161: 189–195.

22. Zhang D, Al-Hendy M, Richard-Davis G, Montgomery-Rice V, Rajaratnam V, Al-Hendy A. Antiproliferative and proapoptotic effects of epigallocatechin gallate on human leiomyoma cells. Fertility and sterility. 2010;94: 1887–1893.

23. Oyeyemi A.O, Oyeyemi R. B, Molehin O. R. Evaluation of Some Mineral Elements and Antioxidant Status in Fibroid Patients in Ado Ekiti, Nigeria. Journal of Research in Pharmaceutical Science. 2016;3: 1–3.

24. Pejić S, Kasapović J, Todorović A, Stojiljković V, Pajović SB. Lipid peroxidation and antioxidant status in blood of patients with uterine myoma, endometrial polypus, hyperplastic and malignant endometrium. Biol Res. 2006;39: 619–629.

25. Feairheller DL, Park J, Sturgeon KM, Williamson ST, Diaz KM, Veerabhadrappa P, et al. Racial Differences in Oxidative Stress and Inflammation: In Vitro and In Vivo. Clin Transl Sci. 2011;4: 32–37.

26. Pepys MB, Hirschfield GM. C-reactive protein: a critical update. The Journal of clinical investigation. 2003;111: 1805–1812.

27. Fletcher NM, Abusamaan MS, Memaj I, Saed MG, Al-Hendy A, Diamond MP, et al. Oxidative stress: a key regulator of leiomyoma cell survival. Fertility and Sterility. 2017;107: 1387–1394.

28. Antolovich M, Prenzler PD, Patsalides E, McDonald S, Robards K. Methods for testing antioxidant activity. Analyst. 2002;127: 183–198.

